# Impact of Epstein-Barr virus co-infection on natural acquired *Plasmodium vivax* antibody response

**DOI:** 10.1101/2022.03.08.22272037

**Authors:** Michelle H F Dias, Luiz F F Guimarães, Matheus G Barcelos, Eduardo U M Moreira, Maria F A do Nascimento, Taís N de Souza, Camilla V Pires, Talita A F Monteiro, Jaap M Middeldorp, Irene S Soares, Francis B Ntumngia, John H Adams, Flora S Kano, Luzia H Carvalho

## Abstract

**Background:** The simultaneous infection of *Plasmodium falciparum* and Epstein-Barr virus (EBV) could promote the development of the aggressive endemic Burkitt’s Lymphoma (eBL) in children living in *P. falciparum* holoendemic areas. While it is well-established that eBL is not related to other human malaria parasites, the impact of EBV infection on the generation of human malaria immunity remains largely unexplored. Considering that this highly prevalent herpesvirus establishes a lifelong persistent infection on B-cells with possible influence on malaria immunity, we hypothesized that EBV co-infection could have impact on the naturally acquired antibody responses to *P. vivax*, the most widespread human malaria parasite.

**Methodology/Principal Findings:** The study design involved three cross-sectional surveys at six-month intervals (baseline, 6 and 12 months) among long-term *P. vivax* exposed adults living in the Amazon rainforest. The approach focused on a group of malaria-exposed individuals whose EBV-DNA (amplification of *balf-5* gene) was persistently detected in the peripheral blood (PersV_DNA_, n=27), and an age-matched malaria-exposed group whose EBV-DNA could never be detected during the follow-up (NegV_DNA_, n=29). During the follow-up period, the serological detection of EBV antibodies to lytic/ latent viral antigens showed that IgG antibodies to viral capsid antigen (VCA-p18) were significantly different between groups (PersV_DNA_ > NegV_DNA_). A panel of blood-stage *P. vivax* antigens covering a wide range of immunogenicity confirmed that in general PersV_DNA_ group showed low levels of antibodies as compared with NegV_DNA_. Interestingly, more significant differences were observed to a novel DBPII immunogen, named DEKnull-2, which has been associated with long-term neutralizing antibody response. Differences between groups were less pronounced with blood-stage antigens (such as MSP1-19) whose levels can fluctuate according to malaria transmission.

**Conclusions/Significance:** In a proof-of-concept study we provide evidence that a persistent detection of EBV DNA in peripheral blood of adults in a *P. vivax* semi-immune population may impact the long-term immune response to major malaria vaccine candidates.

**Author Summary:** In the Amazon rain forest, both *Plasmodium vivax* and Epstein-Barr virus (EBV) infections are common, yet the Burkitt’s lymphoma (BL) is rare, despite an association between endemic BL with chronic *P. falciparum* exposure. Nevertheless, the influence of EBV infection on malaria immunity remains undetermined. Here, we investigated for the first time whether continuous detection of EBV DNA in the peripheral blood of adults exposed to *P. vivax* could impact the antibody response to blood-stage malaria vaccine candidates. The methodological approach involved 12-month follow-up among *P. vivax*-exposed Amazonian classified as persistent EBV-DNA carriers (PersV_DNA_) and an age-matched group with no viral DNA detection (NegV_DNA_); groups were further differentiated based on profile of viral antibodies (mainly IgG VCA-p18). Collectively, our findings demonstrated that antibody levels against *P. vivax* antigens were in general lower in the PersV_DNA_ group as compared with NegV_DNA_. More significant differences were observed to a novel vaccine candidate (DEKnull-2) whose antibody response were previously associated with broadly neutralizing *P. vivax* antibodies. Differences between groups were less pronounced with *P. vivax* antigens associated with seasonal changes in the antibody responses. In this conceptual study, we provide evidence that long-term detection of EBV in peripheral blood may impact immune response to major malaria vaccine candidates.

## Introduction

The impact of malaria infection is greater on populations living in the poorest regions of the globe, where sanitary conditions are precarious, leaving the population subject to possible co-infections with other infectious agents, including parasites [1, 2], bacteria [3, 4] and viruses [5, 6]. Consequently, in malaria endemic areas, the simultaneous infection with multiple pathogens has implications for understanding the development of protective immunity as well as the efficacy of antimalarial vaccines [7, 8]. Common co-infections include herpes viruses as most vertebrates are infected with one or more types that remain for the rest of their lives [9].

The severe consequence of co-infection involving holoendemic *Plasmodium falciparum* exposure and the Epstein-Barr virus (EBV) - a gammaherpes virus that infects B cells and maintains latency throughout the individuals’ lifetime [10, 11] - is well established, as the interaction could promote the development of endemic Burkitt’s Lymphoma (eBL) [12, 13]. In Sub-Saharan Africa, eBL is the most lethal of childhood cancers, with the highest prevalence in children aged 5– 9 years old who are chronically exposed to *P. falciparum* malaria (revised by [14, 15]). Malaria appears to play multiple roles in eBL etiology, including the expansion of latently infected B-cells and the likelihood of c-myc translocation that is a hallmark of BL tumors [16-19]. Of relevance, the aggressive eBL childhood cancer seems to be exclusively linked to *P. falciparum* exposure, but not to other human malaria parasites [20].

Despite the compelling evidence indicating a role for *P. falciparum* in impaired immune responses that control EBV infection [21-23], the impact of acute EBV infection on the generation of anti-malarial immunity is uncertain [24]. Notwithstanding, rodent models of EBV demonstrate that it is possible for a primary gammaherpes virus infection to negatively modulate the generation of antimalarial immunity [7]; an outcome that was correlated with a defect on the generation of humoral immunity to a secondary malaria infection. Evidence of the immune suppressive nature of an acute EBV infection on the development of malaria immunity has also been suggested in experimentally marmosets co-infected with EBV and the quartan malaria *P. brasilianum* [25].

Considering the B-cell compartment as the primary niche for EBV persistence [11] and that humoral malaria immunity may be altered during EBV co-infection [7], we hypothesized here that EBV co-infection could impact on the naturally acquired antibody responses to *P. vivax*, the most geographically widespread human malaria parasite [26]. Taken in to account the rapid spread of *P. vivax* drug-resistant strains [27], and the potential for relapse, progress towards the development of a *P. vivax* vaccine is critical (reviewed in [28]). In this perspective, studying *P. vivax* immunity in the context of EBV co-infections can enhance our understanding of malaria-protective immunity and progress towards the design of next-generation malaria vaccines. Here, we took advantage of a longitudinal follow-up study previously carried out in the Amazon rainforest, where different profiles of *P. vivax* IgG responders were identified [29-31]. To investigate whether a persistent EBV infection could interfere with the profile of *P. vivax* antibody response, we examined in the study population the presence of circulating viral DNA over the time as well as EBV antibody response to lytic viral capsid antigen -VCA, replication activator protein – ZEBRA, early diffuse antigen-EAd, and latent EBV nuclear antigen 1 - EBNA-1.

## Methods

### Area and Study population

The study was carried-out in the agricultural settlement of Rio Pardo (1°46’S—1°54’S, 60°22’W—60°10’W), Presidente Figueiredo municipality, Northeast of Amazonas State in the Brazilian Amazon region. The study site and malaria transmission patterns were described in detail elsewhere [29, 32]. In this area, malaria transmission is considered hypo to mesoendemic, and most residents were natives of the Amazon region. Inhabitants of the settlement live on subsistence farming and fishing along the small streams. In the study area, *P. falciparum* malaria incidence has decreased drastically in recent years, and *P. vivax* is now responsible for all clinical malaria cases reported.

### Study design and cross-sectional surveys

A population-based open cohort study was initiated in November of 2008 and included three cross-sectional surveys carried at six-months interval (baseline, 6 and 12-months), and distributed in periods of high and low malaria transmission (**S1 Fig. 1**), as previously reported [29]. Briefly, (i) interviews were conducted through a structured questionnaire to obtain demographical, epidemiological, and clinical data; (ii) physical examination, including body temperature and spleen/liver size were recorded according to standard clinical protocols; (iii) venous blood was collected for individuals aged five years or older (EDTA, 5 mL); and (iv) examination of Giemsa-stained thick blood smears for the presence of malaria parasites by conventional light microscopy, with *P. vivax* infection confirmed later by a species-specific real-time PCR as described [33]. The geographical location of each dwelling was recorded using a hand-held 12-channel global positioning system (GPS) (Garmin 12XL, Olathe, KS, USA) with a positional accuracy of within 15 m. For the current study, the non-eligible criteria were (i) refusal to sign the informed consent; (ii) children, as clinical immunity is not prevalent in Amazon children [34]; (iii) pregnant women; (iv) any other morbidity that could be traced; and (v) individuals who were unable to be recruited during all three consecutive cross-sectional surveys. Initially, 360 participants were eligible to the current study. The methodological strategy further involved screening these malaria-exposed individuals according to the detection of circulating viral DNA during all cross-sectional surveys (baseline, 6-months, 12-months). For the current study, we focused on two sub-groups of malaria-exposed individuals (i) individuals whose EBV-DNA could be persistently detected from peripheral blood (PersV_DNA_) and (ii) an age-matched subgroup whose viral DNA was undetected throughout the follow-up period (NegV_DNA_) (**Table 1**).

**Table 1.** Demographic, epidemiological, and parasitological data of malaria-exposed individuals whose EBV-DNA could be detected (PersVDNA) or not (NegVDNA) in the peripheral blood during the 12-month follow-up period

|  | PersV<sub>DNA</sub><br>(n = 27) | NegV<sub>DNA</sub><br>(n =29) | p value |
| --- | --- | --- | --- |
| **Characteristic** |  |  |  |
| Age, median, years (IQR) <sup>a</sup> | 38 (22- 54) | 33 (27- 42) | p= 0.1227 |
| Gender ratio, male:female | 1.5/1 | 0.9/1 | p= 0.4357 |
| Years of malaria exposure, median (IQR) <sup>b</sup> | 32 (16-51) | 33 (25-40) | p= 0.7233 |
| Years of residence in Rio Pardo, median (IQR) <sup>c</sup> | 8 (4-10) | 9 (3-14) | p= 0.2326 |
| Riverine population, n (%) <sup>d</sup> | 4 (15%) | 6 (21%) | p= 0.7308 |
| Self-reported malaria episodes, median (IQR) | 5 (2-11) | 11 (3-21) | p= 0.1410 |
| Acute *Plasmodium vivax* infection, n (%) <sup>e</sup> |  |  |  |
| Baseline | 6 (22%) | 6 (21%) |  |
| 6 months | 1 (4%) | 3 (10%) |  |
| 12 months | 4 (15%) | 5 (17%) |  |

<sup>a</sup> IQR = interquartile range
<sup>b</sup> Time living in the endemic area (Brazilian Amazon)
<sup>c</sup> Time living in the agricultural settlement area
<sup>d</sup> Population that living along the Rio Pardo stream.
<sup>e</sup> Asymptomatic malaria infection was detected by conventional light microscopy and real-time PCR

**Fig 1.**
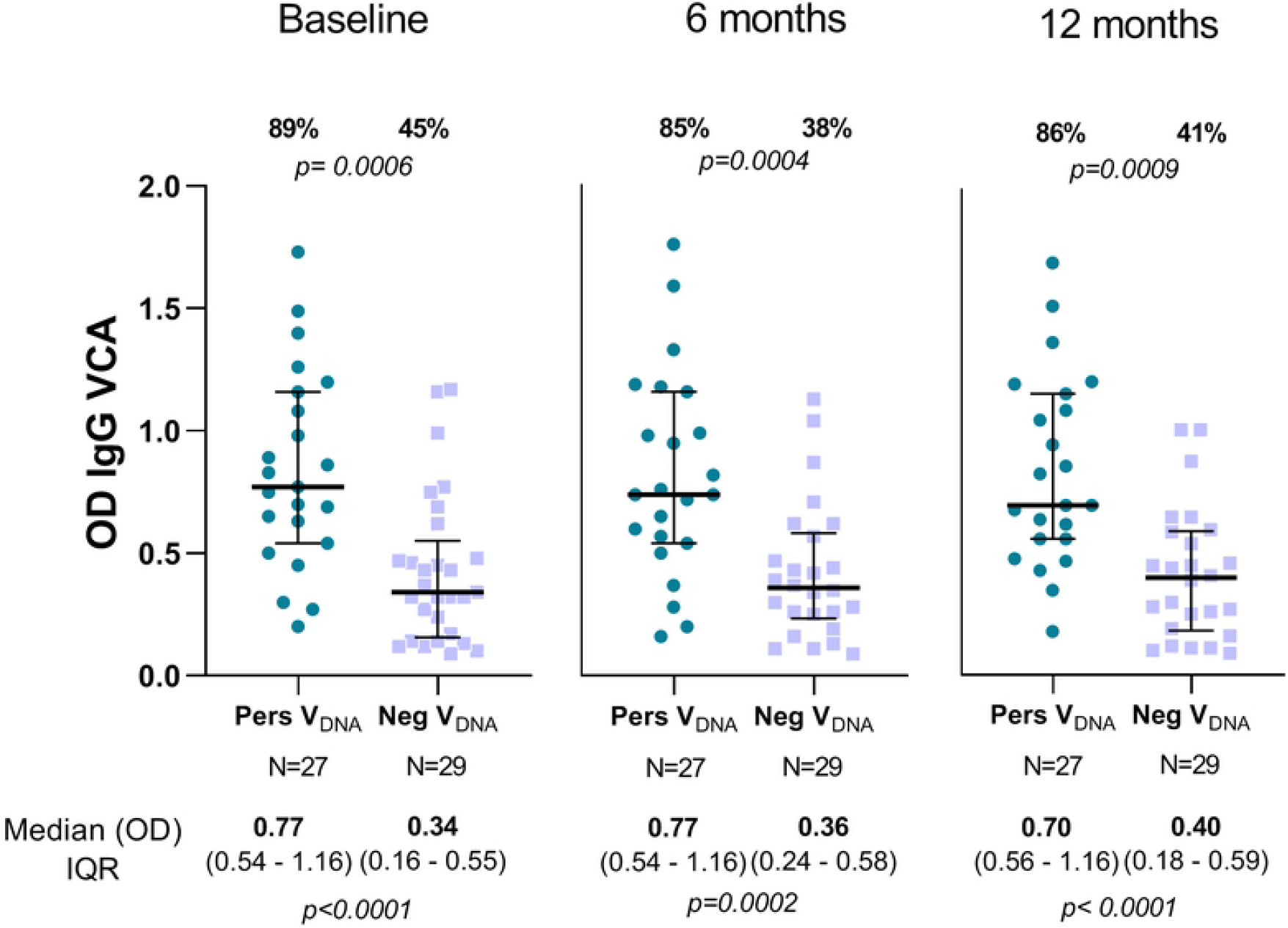
**Profile of IgG antibody response against EBV capsid antigen p18 (VCA-p18) in individuals whose EBV-DNA could be detected (PersV**_**DNA**_**) or not (NegV**_**DNA**_**) over the follow-up period.** For each graphic, individual datapoints were expressed as ELISA absorbance (OD) and shown here as a scatter dot plots with lines showing the median with interquartile range (IQR). Numbers represented in the top and bottom of graphics represent the proportion of responders (%) and median (OD) with IQR values, respectively; p-values for significant difference between groups were included and calculated as described in methods. The proportion of responders was determined by considering an OD> 0.38 as ELISA-positive response (S2 Fig.). All raw data are available in the S1 Table.

The ethical and methodological aspects of this study were approved by the Ethical Committee of Research on Human Beings from the René Rachou Institute (Reports No. 007/2006, No. 07/2009, No.12/2010, No. 26/2013 and CAAE 50522115.7.0000.5091), according to the Resolutions of the Brazilian Council on Health (CNS-196/96 and CNS-466/2012). Formal written consent was obtained from all participants, which was also obtained from the next of kin, caregivers, or guardians on the behalf of child participants.

### Recombinant blood stage *P. vivax* proteins and IgG antibodies detection

***DBPII-related antigens. DBPII-Sal1***, a recombinant Duffy binding protein region II (DBPII) including amino acids 243–573 of the Sal-1 reference strain [35], and recombinant ***DEKnull-2***, an engineered DBPII immunogen [36]. These proteins were expressed as a 39kDa 6xHis fusion protein, properly refolded, as previously described [36, 37]. ***MSP1-19 antigen***. The 19-kDa C-terminal region of the Merozoite Surface Protein-1 of *P. vivax* (MSP1-19), which represents amino acids 1616–1704 of the full-length MSP-1 polypeptide, has been described elsewhere [38]. ***AMA-1 antigen***. The ectodomain of *P. vivax* Apical Membrane Antigen-1 (AMA-1, encompassing amino acids 43 to 487, were produced as previously described [39]. To enable purification, MSP1-19 and AMA-1 constructs were also produced as carboxyl-terminal 6xHis-tag fusion proteins from *Escherichia coli* and *Pichia pastoris*, respectively. ***Conventional Enzyme-Linked Immunoassays (ELISA) for P. vivax IgG antibodies*** was carried out using *P. vivax* blood-stage recombinant proteins as previously described [30], with serum samples at a dilution of 1:100. Recombinant proteins were used at a final concentration of either 3 μg/mL (DBPII and DEKnull-2) or 1 μg/mL (MSP1-19 and AMA-1). For each protein, the results were expressed as ELISA reactivity index (RI), calculated as the ratio of the mean optical density (OD at 492 nm) of each sample to the mean OD plus three standard deviations of samples from 30 unexposed volunteers. Values of RI > 1.0 were considered seropositive.

### EBV antigens and serostatus

EBV-specific antibodies were detected using 4 synthetic peptides covering immunodominant epitopes of the viral capsid antigen P18 (VCA-p18 [BFRF3]), EBV nuclear antigen 1 (EBNA1 [BKRF1]), early diffuse antigen complex (EAd-p45/52 [BMRF1]) and BZLF1-encoded replication activator protein of EBV (Zebra [BZLF1]) [40, 41]. All synthetic peptides were kindly provided by Dr. J. M. Middeldorp (VU University Medical Center, Amsterdam, Netherland). For the assessment of the levels of antibodies to lytic (VCA-p18, EAd and Zebra) and latent EBV antigens (EBNA1), we used synthetic peptide-based ELISA assays as described [42, 43]. Briefly, each peptide was used at final concentration of 1 μg/mL with plasma samples diluted 1:100. IgM (VCA, Zebra and EAd) and IgG (VCA and EBNA-1) reactivities were determined using commercial anti-human IgM and IgG secondary antibodies conjugated to horseradish to peroxidase (HRP) (Sigma-Aldrich). For each peptide, EBV seronegative (n=5) and positive controls (n=8) were used on each experiment. Receiver-operating characteristic (ROC) curves was used to determine optimal cutoff points for each peptide (**S2 Fig**.). Based on the area under the ROC curve (AUC) the follow ELISA’s cutoff were established: (i) 0.38 for VCA IgG (82% sensitivity; 83% specificity), (ii) 0.20 for EBNA-1 IgG (75% sensitivity; 100% specificity), (iii) 0.43 for VCA IgM (82% sensitivity; 100% specificity), (iv) 0.37 for ZEBRA IgM (90% sensitivity; 100% specificity) and (v) 0.44 for EA-d IgM (85% sensitivity; 100% specificity).

### EBV DNA detection by real-time PCR

The PCR primers for this assay were previously selected in the BALF-5 gene encoding the viral DNA polymerase [44]; the upstream and downstream primer sequences were 5′-CGGAAGCCCTCTGGACTTC-3′ and 5′-CCCTGTTTATCCGATGGAATG-3′, respectively, with a fluorogenic probe (5′-TGTACACGCACGAGAAATGCGCC-3′) with a sequence located between the PCR primers. Detectable DNA from EBV was identified by a real-time PCR assay as previously described [45, 46]. Briefly, DNA from whole blood samples collected in EDTA was extracted using Purogene blood core kit B (Qiagen, Minneapolis, MN, USA). The PCR reaction was performed using a mixture containing 1μL of DNA, 0.2 μM each primer, 0.1 μM fluorogenic probe, and 5 μL of TaqMan Master Mix (PE Applied Biosystems), and the PCR cycle was performed as follows: 2 min at 50 °C, 10 min at 95 °C, and 40 cycles of 15 s at 95 °C and 1 min at 60 °C. The TaqMan Master mix (PE Applied Biosystems) was used for all reactions. For all PCR analysis, water was used as negative control, and B958 and P3HR1 viral DNA were used as positive controls. The B958 and P3HR1 viral strains were kindly provided by Dr. Talita A. F. Monteiro (Federal University of Pará, PA, Brazil) and were described elsewhere [47]. Samples were defined as negative if the C_T_ values exceeded 40 cycles.

### Statistical analysis

A database was created using Epidata software (http://www.epidata.dk). The graphics and the statistical analysis were performed using GraphPad Prism version 9.1.2 - GraphPad Software, La Jolla California USA. Receiver operating characteristic curves (ROC) analysis was used to determine optimal Cut-off values for EBV peptides in ELISA assays; Kruska-Wallis or Mann-Whitney test were used to compare median values; chi-square test or Fisher’s exact test were used to compare proportions; and correlations between *P. vivax* and EBV antibody responses were examined by Pearson’s or Spearman’s matrices. In all analysis, a significance level of 5 % was considered, i.e., values of *P* < 0.05.

## Results

### Characteristic of *P. vivax* malaria-exposed groups

At the time of the first cross-sectional survey, 123 (34%) out of 360 Amazonian individuals initially eligible for the study had detectable EBV-DNA in the peripheral blood. Further surveys (6- and 12-month later) identified 27 out of 123 individuals as persistent viral DNA carriers, i.e., individuals whose EBV-DNA was amplified (*balf-5* gene) from the peripheral blood during all follow-up period (PersV_DNA_). In parallel, we selected a group of 29 age-matched malaria-exposed individuals with no detectable viral DNA in the peripheral blood (NegV_DNA_) (**Table 1**). Demographic, parasitological, and epidemiological variables were comparable between groups. Accordingly, most individuals were adults with similar proportion of male: female, and their age basically corresponding to their lifetime exposure to malaria in the Amazon area (medians of 32 and 33 years for PersV_DNA_ and NegV_DNA_, respectively). In this long-term malaria exposed individuals, few acute malaria infections were detected during the follow-up period (all *P. vivax*, as detected by microscopy and/or species-specific PCR assay) (**Table 1**).

### EBV serostatus of *P. vivax* malaria-exposed groups

In these immunocompetent adults, all individuals were seropositive for at least one EBV peptide during the follow-up period (**S3 Fig.)**. Despite of that, individuals categorized as PersV_DNA_ had a much broader EBV antibody response than NegV_DNA_ group; specifically, while 42% of EBV-DNA carriers respond to all five EBV-ELISA markers, only 15%of NegV_DNA_ group responded to these markers (chi-square test = 4.276; *p= 0*.*038*). A similar profile of response was detected over the time (**S3 Fig**.) From the panel of 4 EBV antigens, only IgG response to the lytic antigen VCA-p18 showed a clear differentiation between PersV_DNA_ and NegV_DNA_ groups (**Figure 1**). At enrollment, while 89% of PersV_DNA_ had a positive IgG antibody response to VCA-p18, only 45% of NegV_DNA_ had IgG VCA-p18 (chi-square test; *p= 0*.*0006*). Of interest, the difference in response between groups remained constant throughout the follow-up period. The levels of antibodies to VCA-p18 were also significantly different between groups (Mann-Whitney test), with levels ranging from 0.70 to 0.77 for PersV_DNA_ and from 0.34 to 0.40 for NegV_DNA_ groups **(Figure 1)**.

Although IgM antibodies to EBV lytic antigens (VCA-p18, EAd-p45/52 and Zebra) did not show any differences between the study groups, at the individual level, a positive IgM response for one EBV antigen was related to positivity for the others (Fig. **S4**); of note, this IgM response profile was sustained throughout the study period. Further, we compared the pattern of humoral antibody responses to all EBV antigens between PersV_DNA_ and NegV_DNA_ (**Figure 2**). In the PersV_DNA_ group, there was a tendency of positive correlation between EBV-specific antibodies, specially to IgM antibodies. On the contrary, NegV_DNA_ group was characterized by a predominance of negative antibody correlations to several EBV antigens.

**Fig 2.**
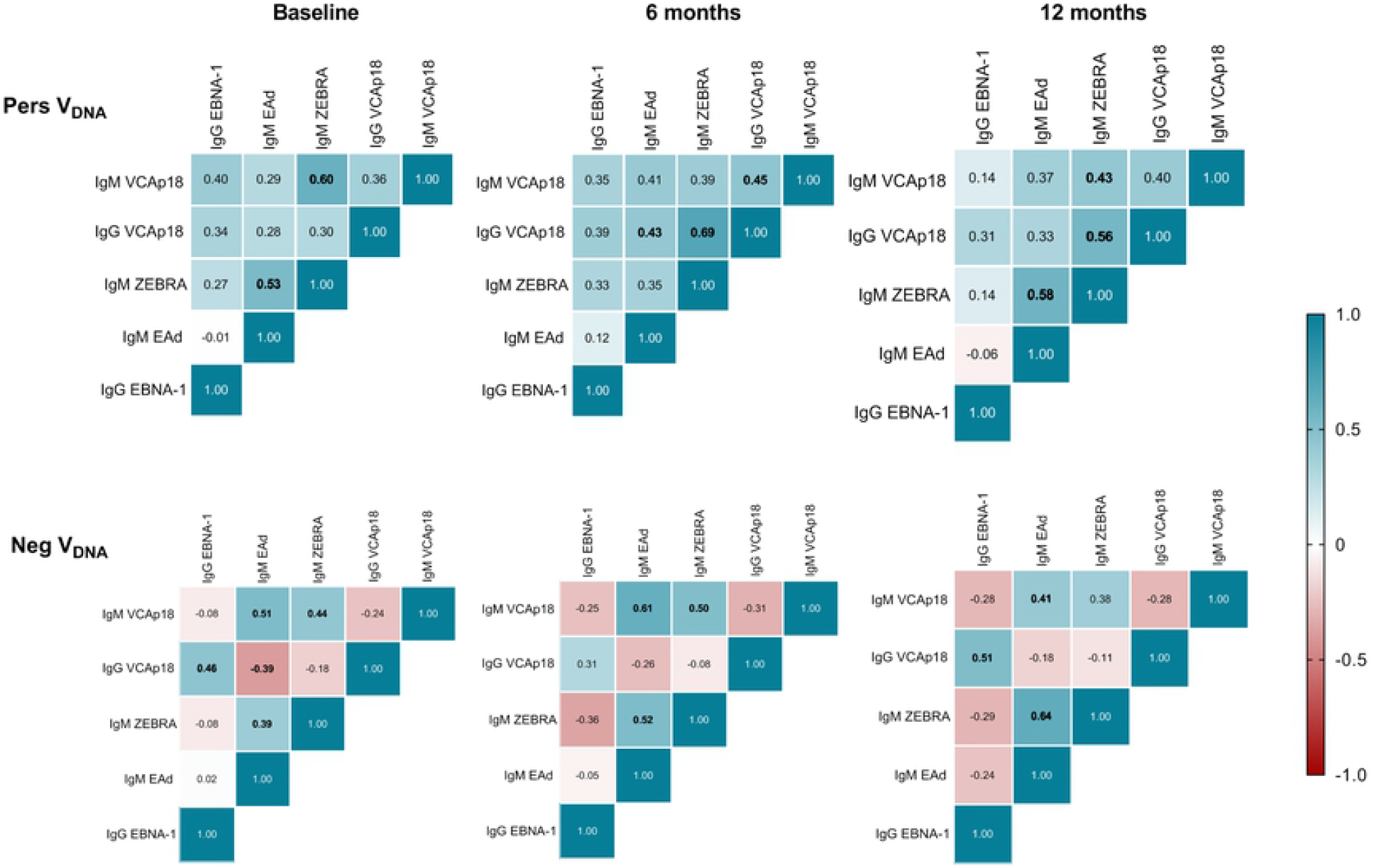
**Pairwise correlations between EBV antibody levels in individuals whose EBV-DNA could be detected (PersV**_**DNA**_**) or not (NegV**_**DNA**_**) over the follow-up period.** Clustering was based on the Spearman correlation coefficient for assays measuring anti-EBV antibodies in plasma samples [IgM (VCA-p18, Zebra and EAd-p45/52) and IgG (VCA-p18 and EBNA-1)]. Matrix heatmaps were shown for each cross-sectional survey (baseline, 6- and 12-months), with top and bottom panels representing PersV_DNA_ and NegV_DNA_ groups, respectively. Positive correlations shown in blue and negative correlations shown in red, with numbers in bold statistically significant differences.

### *Plasmodium vivax* antigen-specific antibody responses in persistent viral DNA carriers

To investigate whether the continuous detection of EBV-DNA in the peripheral blood would impact the humoral response to *P. vivax* malaria, we evaluated antibodies to leading *P. vivax* blood-stage vaccine candidates (DBPII-related antigens, AMA-1 and MSP1-19). In general, there was no clear difference in the proportion of responders between groups, however, the levels of antibody response to different *P. vivax* antigens were significantly different between PersV_DNA_ and NegV_DNA_. The antibody response to the engineered DBPII immunogen (DEKnull-2) showed a trend towards lower antibody levels in PersV_DNA_ compared with NegV_DNA_ (**Figure 3**). Specifically, while DEKnull-2 serological reactivity index (RI) ranged from 0.59 to 0.95 for PersV_DNA_, RI values ranged from 2.11 to 3 for NegV_DNA_ group. Considering all samples assayed, the frequency of positivity was 40% vs. 61% (chi-square test, *p=0*.*0079*) for PersV_DNA_ and NegV_DNA_ samples, respectively. A similar pattern was observed with the original DBPII protein (Fig. **S5**). For *P. vivax* MSP1-19, the differences between the groups was less pronounced (**Figure 4**), with statistically significant differences observed only during high transmission period at the study baseline (**Figure 4**). Circulating antibodies against AMA-1 showed a similar pattern of response as MSP1-19 (Fig. **S6)**.

**Fig 3.**
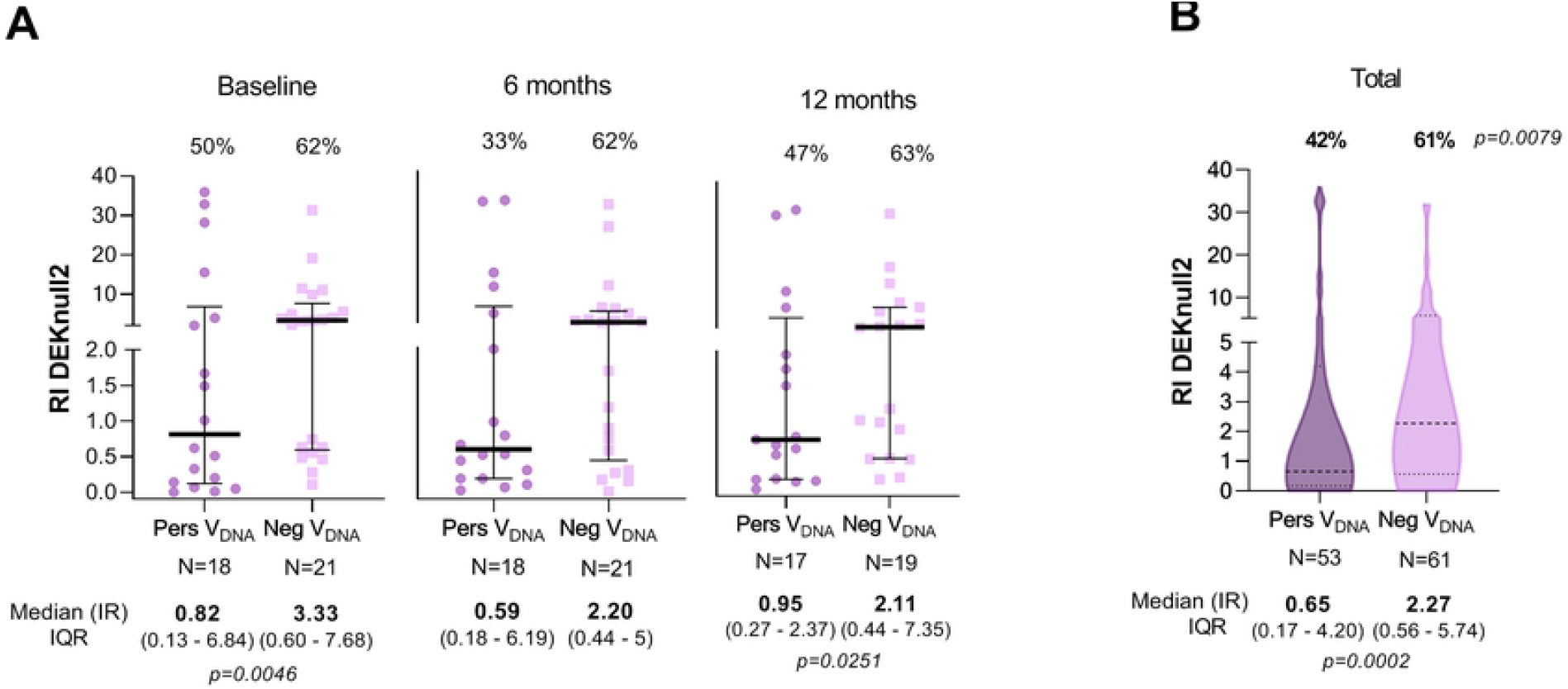
**Profile of IgG antibody response against the surface-engineered DEKnull-2 vaccine of *P. vivax* in individuals with (PersV**_**DNA**_**) or without (NegV**_**DNA**_**) persistent viral DNA over the follow-up period.** In **A**, results are shown by cross-sectional surveys (baseline, 6- and 12-month), with individual datapoints expressed as ELISA Reactivity Index (RI) and shown here as a scatter dot plots with lines showing the median with interquartile range (IQR). In **B**, distribution shape of data (all samples) is represented by violin plots with medians and IQR ranges represented as dotted lines. Numbers in the top and bottom of each graphic represent the proportion of responders (%) and median (RI) with IQR values, respectively; p-values for significant difference between groups were included and calculated as described in methods. Reactivity Index (RI) was calculated as described in methods and RI > 1 corresponded to an ELISA-positive response.

**Fig 4.**
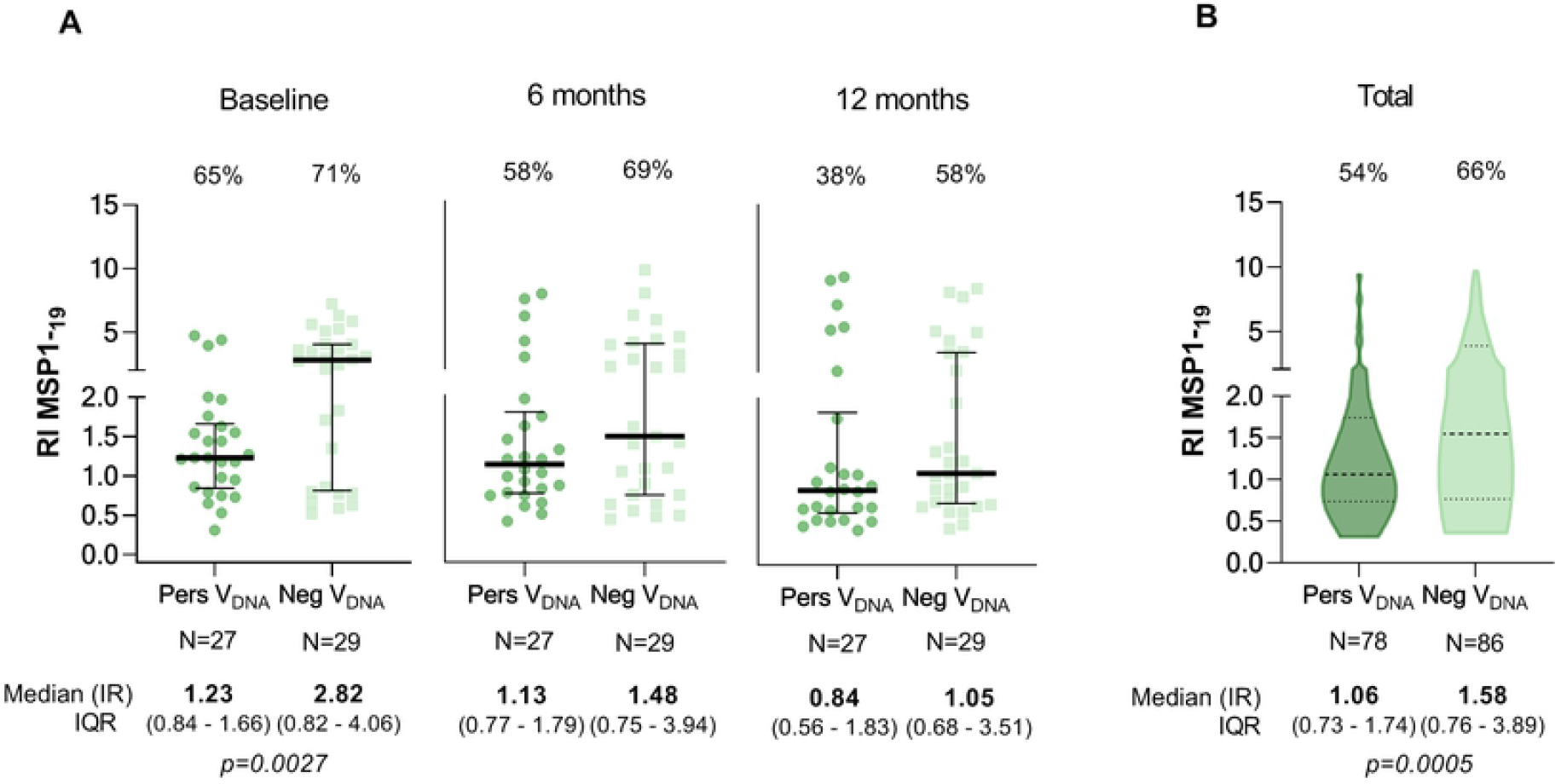
**Profile of IgG antibody response against the 19-kDa C-terminal region of the Merozoite Surface Protein-1 (MSP1-19) in individuals with (PersV**_**DNA**_**) or without (NegV**_**DNA**_**) persistent viral DNA over the follow-up period.** In **A**, results are shown by cross-sectional surveys (baseline, 6- and 12-month), with individual datapoints expressed as ELISA Reactivity Index (RI) and shown here as a scatter dot plots with lines showing the median with interquartile range (IQR). In **B**, distribution shape of data (all samples) is represented by violin plots with medians and IQR ranges represented as dotted lines. Numbers in the top and bottom of each graphic represent the proportion of responders (%) and median (RI) with IQR values, respectively; p-values for significant difference between groups were included and calculated as described in methods. Reactivity Index (RI) was calculated as described in methods and RI > 1 corresponded to an ELISA-positive response.

Regardless, the detection of EBV-DNA in peripheral blood, antibodies against *P. vivax* blood antigens did not correlate with EBV antibody pattern. In both groups, antibody response to *P. vivax* blood-stage antigens showed overall a weak correlation with IgG EBV pattern (sometimes negative associations were also observed) **(Fig. S7)**.

## Discussion

In the Amazon rain forest both malaria and EBV infections are common, yet Burkitt’s lymphoma (BL) is rare [48]; this is not unexpected as endemic BL (eBL) is linked to *P. falciparum* exposure, but not to other malaria parasites [20]. While *P. vivax* malaria is a public health problem in the Amazon, immunological outcomes of prevalent viral co-infections are often neglected [49-51]. Here, we sought to investigate whether a continuous detection of EBV-DNA in peripheral blood of *P. vivax* malaria-exposed adults (PersV_DNA_) could impact antibody response to key *P. vivax* blood-stage vaccine candidates. In this context, it is possible to speculate that a positive PersV_DNA_ status may reflect a reactive EBV persistence with associated deregulated (activated) B-cell function. A 12-month follow-up study demonstrated that levels of *P. vivax*-specific antibodies were in general lower in the PersV_DNA_ group compared with age-matched negative DNA carriers living in the same malaria-endemic village (NegV_DNA_). Interestingly, significant differences in antibody levels were observed against a novel DBPII immunogen, DEKnull-2, that has been associated with stronger, broader, and long-term neutralizing antibody response in the study area [31, 36]. For DEKnull-2, the difference in the magnitude of the antibody response between PersVDNA and NegV_DNA_ groups ranged from two to four-fold, and these differences were maintained throughout the follow-up period. While DEKnull-2 was developed to overcome the inherent DBPII bias towards developing strain-specific immunity associated with poor immunogenicity [52], our methodological approach also included a common DBPII variant (Sal1) circulating in the Amazon area [29]. A similar profile of response was observed for the native Sal1-DBPII variant, in which less reactivity was observed in the PersV_DNA_ group. Taken together, the results suggested that long episodes of EBV-DNA detection may influence the levels of both strain-specific and strain-transcending DBPII immune responses.

With respect to more immunogenic blood stage *P. vivax* antigens, such as MSP1-19 [53], the difference in the magnitude of antibody response between the groups was less pronounced and observed only in samples collected at baseline (PersV_DNA_ < NegV_DNA_), when malaria transmission was more evident in the study area. Although the reasons for the difference in the profile of antibody response between *P. vivax* antigens are not known, we can speculate that local seasonal variation in malaria transmission may have played a role, as antibodies against MSP1-19 /AMA-1 may fluctuate considerably according to malaria transmission [54-56]. In accordance, we previously demonstrated that levels of MSP1-19/AMA-1 (but not DBPII-specific antigens) dropped when malaria transmission was reduced [30]. Considering that our study design involved temporal variation in malaria transmission profile (S1 Fig.), it is reasonable to consider that lowest levels of these antibodies detected at 6 and 12 months of the follow-up (i.e., in the NegV_DNA_ group) may have masked the difference between PersVDNA and NegVDNA groups. Even though our sample size precluded a more robust statistical analysis, the median for MSP1-19 dropped from 3.7 (at enrollment) to 0.8-1.16 (6 and 12 months, respectively), and for AMA-1 from 2.82 to 1.48-1 .05. Future studies should consider the wide range of immunogenicity between P. *vivax* blood stage proteins.

It is also worth mentioning that in this immunocompetent malaria-exposed adult population, we avoid to staging EBV infection based on serology. However, EBV serological profile was only used here as complementary tool to describe the serostatus of PersV_DNA_ versus NegV_DNA_ group. Despite this, PersV_DNA_ had a much broader EBV antibody response than the NegVDNA group, with a significant proportion of viral DNA carriers recognizing all 5 serological markers used here. Considering individual EBV serological markers, only IgG VCA-p18 distinguished PersV_DNA_ and NegV_DNA_ groups during the follow-up study, (85-89% vs. 38-45%, respectively). Interestingly, measuring VCA-IgG antibodies seems to be a best single test to indicate a previous EBV infection [57]. Notwithstanding, in the PersV_DNA_ group, IgG-VCA antibodies tended to be associated with IgM antibodies to lytic antigens (VCA-p18, EAd-p45/52 and Zebra). Intriguingly, EBV-IgM responses were relatively stable during the follow-up period (in both groups for all EBV antigens). Although the reasons for these findings are unclear, concerns have been raised about possible cross-reactivities of EBV-IgM antibodies with other antigenically related viral infections [58-60], such as CMV that is prevalent in the Amazon area [61]. Although IgM cross-reactivity could not be ruled out, the tendency of correlation with EBV-DNA may reflect constant EBV activation-triggering of B-cells; in fact, EBV persistently infecting naive or anti-EBV B-cells leading to increased IgM levels [62, 63]. Finally, it is important to clarify about the high sensitivity and specificity of the EBV-peptides used in our ELISA assays. According, in a group of 34 Amazonian children (median age 9 years, IQR 8-11) whose plasma samples were screened for anti-EBV antibodies using the same peptide-based ELISA protocol, we found that VCA-IgM and Zebra-IgM antibodies decreased over time while VCA-IgG and EBNA-IgG increased within same period of time (12-month follow-up), and remained positive throughout the observation period (**Fig.S8**). These results confirmed the typical pattern of seroconversion of young children with low socioeconomic status [64], correlating with an early EBV seroconversion for Latin America population [65]. Consequently, the reason for the persistence of the IgM response in our adult cohort merits further investigations.

As expected, few malaria cases were detected in our malaria semi-immune study population, which precluded our ability to evaluate an association between acute *P. vivax* infection and persistent detection of EBV-DNA. Although the current study was not designed to investigate whether EBV can change the course of an acute *P. vivax* malaria infection, scant data from Indonesia suggest that whereas EBV-DNA levels were significantly elevated in high parasitemic *P. vivax* individuals, EBV-DNA levels were not related to age, gender, or malaria symptoms [66]. To properly address the influence of the persistence of EBV-DNA detection in the course of vivax malaria, a non-immune symptomatic *P. vivax* population should be investigated that was out of the scope of the current MS.

Our study has limitations that should be considered when interpreting the results. First, a relatively small number of malaria-exposed residents were eligible to participate in the study, which may have underpowered some statistical analyses. Specifically, the 12-month cohort study demonstrated that only 27 out of 123 individuals presented persistent detection of EBV-DNA in peripheral blood. Despite this limitation, we are confident about the robustness of the study design, which allowed us to demonstrate for the first time that antibody levels to different *P. vivax* antigens were significantly lower in subjects with persistent EBV “DNAmia”. As the main host cell of the EBV is the human B cell (reviewed by [67]), ongoing experiments are in progress to evaluate on the role of a persistent detection of EBV-DNA in the long-term *P. vivax*-specific B cell response. In this proof of concept study, we provide evidence that a persistent detection of EBV in peripheral blood of an adult *P. vivax* semi-immune population may impact the long-term malaria immune response to major malaria vaccine candidates.

## Data Availability

All data produced in the present work are contained in the manuscript

## Acknowledgments

We thank the inhabitants of Rio Pardo for enthusiastic participation in the study; the local malaria control team in Presidente Fiqueiredo for their logistic support; the units of Fundação Oswaldo Cruz in Manaus, AM (Fiocruz Amazonia), and Belo Horizonte, MG (Fiocruz Minas), for overall support. The Program for Technological Development in Tools for Health-PDTIS-FIOCRUZ for use of its facilities at René Rachou Institute (Real-Time PCR Facility; RPT09D). The Coordination for the Improvement of Higher Education Personnel (CAPES) and the Program for Institutional Internationalization of CAPES-PrInt/ FIOCRUZ is also acknowledged.

## Supporting Information Citations

**S1 Fig. Monthly-time series of malaria cases in the agricultural settlement of Rio Pardo (Amazonas, Brazil) during the study period, 2008–2009**. The current study included three cross-sectional surveys at six-month intervals (Baseline, Bs; 6- and 12-months latter). Malaria cases were based on results of conventional microscopy provided by the National Malaria Surveillance System Registry (SIVEP-Malaria), with cases of *P. falciparum* (light blue) and *P. vivax* (dark blue) plotted per month.

**S2 Fig. Two-graph receiver operating characteristic curves (TG-ROC) to EBV proteins**. For ELISA-detected antibodies [IgM (VCA-p18, Zebra and EAd-p45/52) and IgG (VCA-p18 and EBNA-1)], the best cutoff values was determined through sensibility and specificity calculated by TG-ROC curves in GraphPad Prism 9.2, as described in material and methods.

**S3 Fig. Frequency of anti-EBV antibodies according to the number of serological markers recognized by individuals whose EBV-DNA could be detected (PersV**_**DNA**_**) or not (NegV**_**DNA**_**) over the follow-up period**. For each group, results were presented as the proportion of responders for one (1), two (2), three (3), four (4) or five (5) EBV serological markers [IgM (VCA-p18, Zebra and EAd-p45/52) and IgG (VCA-p18 and EBNA-1)]. All raw data are available in the S1 Table.

**S4 Fig. Individual antibody response to Epstein-Barr virus (EBV) peptides (VCA-p18, ZEBRA, EAd-p45/52 and EBNA-1) during the cross-sectional surveys**. Heatmaps illustrated individual antibody response of each malaria-exposed individuals classified according to the detection (PersV_DNA_) or not (NegV_DNA_) of EBV-DNA over the follow-up period. According to EBV antibody response, individuals were categorized as non-responder (negative) or responders (stratified as low, medium or high, according to EBV antibody reactivity). The missing values are recorded as blank spaces.

**S5 Fig. Profile of IgG antibody response against *P. vivax* r Duffy Binding Protein region II (DBPII) in individuals with (PersV**_**DNA**_**) or without (NegV**_**DNA**_**) persistent viral DNA over the follow-up period**. In **A**, results are shown by cross-sectional surveys (baseline, 6- and 12-month), with individual datapoints expressed as ELISA Reactivity Index (RI) and shown here as a scatter dot plots with lines showing the median with interquartile range (IQR). In **B**, distribution shape of data (all samples) is represented by violin plots with medians and IQR ranges represented as dotted lines. Numbers in the top and bottom of each graphic represent the proportion of responders (%) and median (RI) with IQR values, respectively; p-values for significant difference between groups were included and calculated as described in methods. Reactivity Index (RI) was calculated as described in methods and RI > 1 corresponded to an ELISA-positive response.

**S6 Fig. Profile of IgG antibody response against *P. vivax* Apical Membrane Antigen-1 (AMA-1), in individuals with (PersV**_**DNA**_**) or without (NegV**_**DNA**_**) persistent viral DNA over the follow-up period**. In **A**, results are shown by cross-sectional surveys (baseline, 6- and 12-month), with individual datapoints expressed as ELISA Reactivity Index (RI) and shown here as a scatter dot plots with lines showing the median with interquartile range (IQR). In **B**, distribution shape of data (all samples) is represented by violin plots with medians and IQR ranges represented as dotted lines. Numbers in the top and bottom of each graphic represent the proportion of responders (%) and median (RI) with IQR values, respectively; p-values for significant difference between groups were included and calculated as described in methods. Reactivity Index (RI) was calculated as described in methods and RI > 1 corresponded to an ELISA-positive response.

**S7 Fig. Pairwise correlations between EBV and *P. vivax* antibody levels in individuals with (PersV**_**DNA**_**) or without (NegV**_**DNA**_**) persistent viral DNA during the cross-sectional surveys**. Clustering was based on the Spearman correlation coefficient for assays measuring anti-EBV antibodies in serum. Matrix heatmaps were shown for each cross-sectional survey (baseline, 6- and 12-months), with top and bottom panels representing Pers V_DNA_ an Neg V_DNA_ groups, respectively. Positive correlations shown in blue and negative correlations shown in orange, with numbers in bold statistically significant differences.

**S8 Fig. IgM and IgG antibody response against Epstein-Barr virus peptides in Amazonian children over time**. For each EBV-peptide (VCA-p18; Zebra, EAd-p45/52 and EBNA-1) antibody response was represented by frequency of responders (bar) and magnitude of response (optical density-OD median, lines). The results represented three cross-sectional surveys carried-out at 6-month intervals, i.e., at enrollment (0m) and six (6m) and 12-month latter (12m). ELISA assays were carried out as described in the methods.

## References

1. Brooker S, Akhwale W, Pullan R, Estambale B, Clarke SE, Snow RW, et al. Epidemiology of plasmodium-helminth co-infection in Africa: populations at risk, potential impact on anemia, and prospects for combining control. Am J Trop Med Hyg. 2007;77(6 Suppl):88–98. Epub 2008/01/31. PubMed PMID: 18165479; PubMed Central PMCID: PMCPMC2637949.

2. Afolabi MO, Ale BM, Dabira ED, Agbla SC, Bustinduy AL, Ndiaye JLA, et al. Malaria and helminth co-infections in children living in endemic countries: A systematic review with meta-analysis. PLoS Negl Trop Dis. 2021;15(2):e0009138. Epub 2021/02/19. doi: 10.1371/journal.pntd.0009138. PubMed PMID: 33600494; PubMed Central PMCID: PMCPMC7924789.

3. Mooney JP, Galloway LJ, Riley EM. Malaria, anemia, and invasive bacterial disease: A neutrophil problem? J Leukoc Biol. 2019;105(4):645–55. Epub 2018/12/21. doi: 10.1002/JLB.3RI1018-400R. PubMed PMID: 30570786; PubMed Central PMCID: PMCPMC6487965.

4. Nyirenda TS, Mandala WL, Gordon MA, Mastroeni P. Immunological bases of increased susceptibility to invasive nontyphoidal Salmonella infection in children with malaria and anaemia. Microbes Infect. 2018;20(9-10):589–98. Epub 2017/12/19. doi: 10.1016/j.micinf.2017.11.014. PubMed PMID: 29248635; PubMed Central PMCID: PMCPMC6250906.

5. Kotepui KU, Kotepui M. Prevalence of and risk factors for Plasmodium spp. co-infection with hepatitis B virus: a systematic review and meta-analysis. Malar J. 2020;19(1):368. Epub 2020/10/17. doi: 10.1186/s12936-020-03428-w. PubMed PMID: 33059662; PubMed Central PMCID: PMCPMC7560023.

6. Roberds A, Ferraro E, Luckhart S, Stewart VA. HIV-1 Impact on Malaria Transmission: A Complex and Relevant Global Health Concern. Front Cell Infect Microbiol. 2021;11:656938. Epub 2021/04/30. doi: 10.3389/fcimb.2021.656938. PubMed PMID: 33912477; PubMed Central PMCID: PMCPMC8071860.

7. Matar CG, Jacobs NT, Speck SH, Lamb TJ, Moormann AM. Does EBV alter the pathogenesis of malaria? Parasite Immunol. 2015;37(9):433–45. Epub 2015/06/30. doi: 10.1111/pim.12212. PubMed PMID: 26121587.

8. Falanga YT, Frascoli M, Kaymaz Y, Forconi C, Ong’echa JM, Bailey JA, et al. High pathogen burden in childhood promotes the development of unconventional innate-like CD8+ T cells. JCI Insight. 2017;2(15). Epub 2017/08/05. doi: 10.1172/jci.insight.93814. PubMed PMID: 28768916; PubMed Central PMCID: PMCPMC5543908.

9. Sehrawat S, Kumar D, Rouse BT. Herpesviruses: Harmonious Pathogens but Relevant Cofactors in Other Diseases? Front Cell Infect Microbiol. 2018;8:177. Epub 2018/06/12. doi: 10.3389/fcimb.2018.00177. PubMed PMID: 29888215; PubMed Central PMCID: PMCPMC5981231.

10. Young LS, Rickinson AB. Epstein-Barr virus: 40 years on. Nat Rev Cancer. 2004;4(10):757–68. Epub 2004/10/29. doi: 10.1038/nrc1452. PubMed PMID: 15510157.

11. Thorley-Lawson DA. EBV Persistence--Introducing the Virus. Curr Top Microbiol Immunol. 2015;390(Pt 1):151–209. Epub 2015/10/02. doi: 10.1007/978-3-319-22822-8_8. PubMed PMID: 26424647; PubMed Central PMCID: PMCPMC5125397.

12. Epstein MA, Achong BG, Barr YM. Virus Particles in Cultured Lymphoblasts from Burkitt’s Lymphoma. Lancet. 1964;1(7335):702–3. Epub 1964/03/28. doi: 10.1016/s0140-6736(64)91524-7. PubMed PMID: 14107961.

13. Kennedy G, Komano J, Sugden B. Epstein-Barr virus provides a survival factor to Burkitt’s lymphomas. Proc Natl Acad Sci U S A. 2003;100(24):14269–74. Epub 2003/11/07. doi: 10.1073/pnas.2336099100. PubMed PMID: 14603034; PubMed Central PMCID: PMCPMC283581.

14. Moormann AM, Bailey JA. Malaria - how this parasitic infection aids and abets EBV-associated Burkitt lymphomagenesis. Curr Opin Virol. 2016;20:78–84. Epub 2016/10/01. doi: 10.1016/j.coviro.2016.09.006. PubMed PMID: 27689909; PubMed Central PMCID: PMCPMC5102755.

15. Jayasooriya S, Hislop A, Peng Y, Croom-Carter D, Jankey Y, Bell A, et al. Revisiting the effect of acute P. falciparum malaria on Epstein-Barr virus: host balance in the setting of reduced malaria endemicity. PLoS One. 2012;7(2):e31142. Epub 2012/02/22. doi: 10.1371/journal.pone.0031142. PubMed PMID: 22347443; PubMed Central PMCID: PMCPMC3275582.

16. Torgbor C, Awuah P, Deitsch K, Kalantari P, Duca KA, Thorley-Lawson DA. A multifactorial role for P. falciparum malaria in endemic Burkitt’s lymphoma pathogenesis. PLoS Pathog. 2014;10(5):e1004170. Epub 2014/05/31. doi: 10.1371/journal.ppat.1004170. PubMed PMID: 24874410; PubMed Central PMCID: PMCPMC4038605.

17. Robbiani DF, Deroubaix S, Feldhahn N, Oliveira TY, Callen E, Wang Q, et al. Plasmodium Infection Promotes Genomic Instability and AID-Dependent B Cell Lymphoma. Cell. 2015;162(4):727–37. Epub 2015/08/16. doi: 10.1016/j.cell.2015.07.019. PubMed PMID: 26276629; PubMed Central PMCID: PMCPMC4538708.

18. Graham BS, Lynch DT. Burkitt Lymphoma. StatPearls. Treasure Island (FL) 2021.

19. Wilmore JR, Asito AS, Wei C, Piriou E, Sumba PO, Sanz I, et al. AID expression in peripheral blood of children living in a malaria holoendemic region is associated with changes in B cell subsets and Epstein-Barr virus. Int J Cancer. 2015;136(6):1371–80. Epub 2014/08/08. doi: 10.1002/ijc.29127. PubMed PMID: 25099163; PubMed Central PMCID: PMCPMC4697832.

20. Quintana MDP, Smith-Togobo C, Moormann A, Hviid L. Endemic Burkitt lymphoma - an aggressive childhood cancer linked to Plasmodium falciparum exposure, but not to exposure to other malaria parasites. APMIS. 2020;128(2):129–35. Epub 2020/03/07. doi: 10.1111/apm.13018. PubMed PMID: 32133709.

21. Moormann AM, Chelimo K, Sumba PO, Tisch DJ, Rochford R, Kazura JW. Exposure to holoendemic malaria results in suppression of Epstein-Barr virus-specific T cell immunosurveillance in Kenyan children. J Infect Dis. 2007;195(6):799–808. Epub 2007/02/15. doi: 10.1086/511984. PubMed PMID: 17299709.

22. Moss DJ, Burrows SR, Castelino DJ, Kane RG, Pope JH, Rickinson AB, et al. A comparison of Epstein-Barr virus-specific T-cell immunity in malaria-endemic and - nonendemic regions of Papua New Guinea. Int J Cancer. 1983;31(6):727–32. Epub 1983/06/15. doi: 10.1002/ijc.2910310609. PubMed PMID: 6305850.

23. Chattopadhyay PK, Chelimo K, Embury PB, Mulama DH, Sumba PO, Gostick E, et al. Holoendemic malaria exposure is associated with altered Epstein-Barr virus-specific CD8(+) T-cell differentiation. J Virol. 2013;87(3):1779–88. Epub 2012/11/24. doi: 10.1128/JVI.02158-12. PubMed PMID: 23175378; PubMed Central PMCID: PMCPMC3554182.

24. Derkach A, Otim I, Pfeiffer RM, Onabajo OO, Legason ID, Nabalende H, et al. Associations between IgG reactivity to Plasmodium falciparum erythrocyte membrane protein 1 (PfEMP1) antigens and Burkitt lymphoma in Ghana and Uganda case-control studies. EBioMedicine. 2019;39:358–68. Epub 2018/12/24. doi:10.1016/j.ebiom.2018.12.020. PubMed PMID: 30579868; PubMed Central PMCID: PMCPMC6355394.

25. Wedderburn N, Davies DR, Mitchell GH, Desgranges C, de The G. Glomerulonephritis in common marmosets infected with Plasmodium brasilianum and Epstein-Barr virus. J Infect Dis. 1988;158(4):789–94. Epub 1988/10/01. doi: 10.1093/infdis/158.4.789. PubMed PMID: 2844917.

26. WHO. World malaria report 2020. Report. Genebra: 2000 November 30. Report No.: ISBN 978-92-4-001579-1

27. Ferreira MU, Nobrega de Sousa T, Rangel GW, Johansen IC, Corder RM, Ladeia-Andrade S, et al. Monitoring Plasmodium vivax resistance to antimalarials: Persisting challenges and future directions. Int J Parasitol Drugs Drug Resist. 2020;15:9–24. Epub 2020/12/29. doi: 10.1016/j.ijpddr.2020.12.001. PubMed PMID: 33360105; PubMed Central PMCID: PMCPMC7770540.

28. De SL, Ntumngia FB, Nicholas J, Adams JH. Progress towards the development of a P. vivax vaccine. Expert Rev Vaccines. 2021:1–16. Epub 2021/01/23. doi: 10.1080/14760584.2021.1880898. PubMed PMID: 33481638.

29. Kano FS, Sanchez BA, Sousa TN, Tang ML, Saliba J, Oliveira FM, et al. Plasmodium vivax Duffy binding protein: baseline antibody responses and parasite polymorphisms in a well-consolidated settlement of the Amazon Region. Trop Med Int Health. 2012;17(8):989–1000. Epub 2012/05/31. doi: 10.1111/j.1365-3156.2012.03016.x. PubMed PMID: 22643072.

30. Pires CV, Alves JRS, Lima BAS, Paula RB, Costa HL, Torres LM, et al. Blood-stage Plasmodium vivax antibody dynamics in a low transmission setting: A nine year follow-up study in the Amazon region. PLoS One. 2018;13(11):e0207244. Epub 2018/11/13. doi: 10.1371/journal.pone.0207244. PubMed PMID: 30419071; PubMed Central PMCID: PMCPMC6231651.

31. Medeiros CMP, Moreira EUM, Pires CV, Torres LM, Guimaraes LFF, Alves JRS, et al. Dynamics of IgM and IgG responses to the next generation of engineered Duffy binding protein II immunogen: Strain-specific and strain-transcending immune responses over a nine-year period. PLoS One. 2020;15(5):e0232786. Epub 2020/05/08. doi: 10.1371/journal.pone.0232786. PubMed PMID: 32379804; PubMed Central PMCID: PMCPMC7205269.

32. Kano FS, Souza-Silva FA, Torres LM, Lima BA, Sousa TN, Alves JR, et al. The Presence, Persistence and Functional Properties of Plasmodium vivax Duffy Binding Protein II Antibodies Are Influenced by HLA Class II Allelic Variants. PLoS Negl Trop Dis. 2016;10(12):e0005177. Epub 2016/12/14. doi: 10.1371/journal.pntd.0005177. PubMed PMID: 27959918; PubMed Central PMCID: PMCPMC5154503.

33. Amaral LC, Robortella DR, Guimaraes LFF, Limongi JE, Fontes CJF, Pereira DB, et al. Ribosomal and non-ribosomal PCR targets for the detection of low-density and mixed malaria infections. Malar J. 2019;18(1):154. Epub 2019/05/02. doi: 10.1186/s12936-019-2781-3. PubMed PMID: 31039781; PubMed Central PMCID: PMCPMC6492410.

34. Ladeia-Andrade S, Ferreira MU, de Carvalho ME, Curado I, Coura JR. Age-dependent acquisition of protective immunity to malaria in riverine populations of the Amazon Basin of Brazil. Am J Trop Med Hyg. 2009;80(3):452–9. Epub 2009/03/10. PubMed PMID: 19270298.

35. Fang XD, Kaslow DC, Adams JH, Miller LH. Cloning of the Plasmodium vivax Duffy receptor. Mol Biochem Parasitol. 1991;44(1):125–32. Epub 1991/01/01. doi: 10.1016/0166-6851(91)90228-x. PubMed PMID: 1849231.

36. Ntumngia FB, Pires CV, Barnes SJ, George MT, Thomson-Luque R, Kano FS, et al. An engineered vaccine of the Plasmodium vivax Duffy binding protein enhances induction of broadly neutralizing antibodies. Sci Rep. 2017;7(1):13779. Epub 2017/10/25. doi: 10.1038/s41598-017-13891-2. PubMed PMID: 29062081; PubMed Central PMCID: PMCPMC5653783.

37. Ntumngia FB, Adams JH. Design and immunogenicity of a novel synthetic antigen based on the ligand domain of the Plasmodium vivax duffy binding protein. Clin Vaccine Immunol. 2012;19(1):30–6. Epub 2011/11/26. doi: 10.1128/CVI.05466-11. PubMed PMID: 22116684; PubMed Central PMCID: PMCPMC3255949.

38. Rodrigues MH, Cunha MG, Machado RL, Ferreira OC, Jr., Rodrigues MM, Soares IS. Serological detection of Plasmodium vivax malaria using recombinant proteins corresponding to the 19-kDa C-terminal region of the merozoite surface protein-1. Malar J. 2003;2(1):39. Epub 20031114. doi: 10.1186/1475-2875-2-39. PubMed PMID: 14617378; PubMed Central PMCID: PMCPMC293434.

39. Vicentin EC, Francoso KS, Rocha MV, Iourtov D, Dos Santos FL, Kubrusly FS, et al. Invasion-inhibitory antibodies elicited by immunization with Plasmodium vivax apical membrane antigen-1 expressed in Pichia pastoris yeast. Infect Immun. 2014;82(3):1296–307. Epub 2014/01/01. doi: 10.1128/IAI.01169-13. PubMed PMID: 24379279; PubMed Central PMCID: PMCPMC3958008.

40. Fachiroh J, Paramita DK, Hariwiyanto B, Harijadi A, Dahlia HL, Indrasari SR, et al. Single-assay combination of Epstein-Barr Virus (EBV) EBNA1- and viral capsid antigen-p18-derived synthetic peptides for measuring anti-EBV immunoglobulin G (IgG) and IgA antibody levels in sera from nasopharyngeal carcinoma patients: options for field screening. J Clin Microbiol. 2006;44(4):1459–67. Epub 2006/04/07. doi: 10.1128/JCM.44.4.1459-1467.2006. PubMed PMID: 16597877; PubMed Central PMCID: PMCPMC1448657.

41. Piriou E, Kimmel R, Chelimo K, Middeldorp JM, Odada PS, Ploutz-Snyder R, et al. Serological evidence for long-term Epstein-Barr virus reactivation in children living in a holoendemic malaria region of Kenya. J Med Virol. 2009;81(6):1088–93. Epub 2009/04/22. doi: 10.1002/jmv.21485. PubMed PMID: 19382256; PubMed Central PMCID: PMCPMC3134942.

42. de Sanjose S, Bosch R, Schouten T, Verkuijlen S, Nieters A, Foretova L, et al. Epstein-Barr virus infection and risk of lymphoma: immunoblot analysis of antibody responses against EBV-related proteins in a large series of lymphoma subjects and matched controls. Int J Cancer. 2007;121(8):1806–12. Epub 2007/06/09. doi: 10.1002/ijc.22857. PubMed PMID: 17557295.

43. Ogolla S, Daud, II, Asito AS, Sumba OP, Ouma C, Vulule J, et al. Reduced Transplacental Transfer of a Subset of Epstein-Barr Virus-Specific Antibodies to Neonates of Mothers Infected with Plasmodium falciparum Malaria during Pregnancy. Clin Vaccine Immunol. 2015;22(11):1197–205. Epub 2015/09/18. doi: 10.1128/CVI.00270-15. PubMed PMID: 26376931; PubMed Central PMCID: PMCPMC4622112.

44. Kimura H, Morita M, Yabuta Y, Kuzushima K, Kato K, Kojima S, et al. Quantitative analysis of Epstein-Barr virus load by using a real-time PCR assay. J Clin Microbiol. 1999;37(1):132–6. Epub 1998/12/17. doi: 10.1128/JCM.37.1.132-136.1999. PubMed PMID: 9854077; PubMed Central PMCID: PMCPMC84187.

45. Piriou E, Asito AS, Sumba PO, Fiore N, Middeldorp JM, Moormann AM, et al. Early age at time of primary Epstein-Barr virus infection results in poorly controlled viral infection in infants from Western Kenya: clues to the etiology of endemic Burkitt lymphoma. J Infect Dis. 2012;205(6):906–13. Epub 2012/02/04. doi: 10.1093/infdis/jir872. PubMed PMID: 22301635; PubMed Central PMCID: PMCPMC3282570.

46. Moormann AM, Chelimo K, Sumba OP, Lutzke ML, Ploutz-Snyder R, Newton D, et al. Exposure to holoendemic malaria results in elevated Epstein-Barr virus loads in children. J Infect Dis. 2005;191(8):1233–8. Epub 2005/03/19. doi: 10.1086/428910. PubMed PMID: 15776368.

47. Monteiro TAF, Costa IB, Costa IB, Correa T, Coelho BMR, Silva AES, et al. Genotypes of Epstein-Barr virus (EBV1/EBV2) in individuals with infectious mononucleosis in the metropolitan area of Belem, Brazil, between 2005 and 2016. Braz J Infect Dis. 2020;24(4):322–9. Epub 20200630. doi: 10.1016/j.bjid.2020.06.004. PubMed PMID: 32619403.

48. Ambrus JL, Ambrus CM. Burkitt’s lymphoma. J Med. 1981;12(6):385-413. PubMed PMID: 6274988.

49. Goncalves RM, Lima NF, Ferreira MU. Parasite virulence, co-infections and cytokine balance in malaria. Pathog Glob Health. 2014;108(4):173–8. Epub 20140523. doi: 10.1179/2047773214Y.0000000139. PubMed PMID: 24854175; PubMed Central PMCID: PMCPMC4069333.

50. Cruz LAB, Moraes MOA, Queiroga-Barros MR, Fukutani KF, Barral-Netto M, Andrade BB. Chronic hepatitis B virus infection drives changes in systemic immune activation profile in patients coinfected with Plasmodium vivax malaria. PLoS Negl Trop Dis. 2019;13(6):e0007535. Epub 20190624. doi: 10.1371/journal.pntd.0007535. PubMed PMID: 31233500; PubMed Central PMCID: PMCPMC6611654.

51. Del-Tejo PL, Cubas-Vega N, Caraballo-Guerra C, da Silva BM, da Silva Valente J, Sampaio VS, et al. Should we care about Plasmodium vivax and HIV co-infection? A systematic review and a cases series from the Brazilian Amazon. Malar J. 2021;20(1):13. Epub 20210106. doi: 10.1186/s12936-020-03518-9. PubMed PMID: 33407474; PubMed Central PMCID: PMCPMC7788992.

52. Ceravolo IP, Sanchez BA, Sousa TN, Guerra BM, Soares IS, Braga EM, et al. Naturally acquired inhibitory antibodies to Plasmodium vivax Duffy binding protein are short-lived and allele-specific following a single malaria infection. Clin Exp Immunol. 2009;156(3):502–10. doi: 10.1111/j.1365-2249.2009.03931.x. PubMed PMID: 19438604; PubMed Central PMCID: PMCPMC2691980.

53. Mueller I, Galinski MR, Tsuboi T, Arevalo-Herrera M, Collins WE, King CL. Natural acquisition of immunity to Plasmodium vivax: epidemiological observations and potential targets. Adv Parasitol. 2013;81:77–131. doi: 10.1016/B978-0-12-407826-0.00003-5. PubMed PMID: 23384622.

54. Cunha MG, Silva ES, Sepulveda N, Costa SP, Saboia TC, Guerreiro JF, et al. Serologically defined variations in malaria endemicity in Para state, Brazil. PLoS One. 2014;9(11):e113357. Epub 20141124. doi: 10.1371/journal.pone.0113357. PubMed PMID: 25419900; PubMed Central PMCID: PMCPMC4242530.

55. Yman V, White MT, Rono J, Arca B, Osier FH, Troye-Blomberg M, et al. Antibody acquisition models: A new tool for serological surveillance of malaria transmission intensity. Sci Rep. 2016;6:19472. Epub 20160205. doi: 10.1038/srep19472. PubMed PMID: 26846726; PubMed Central PMCID: PMCPMC4984902.

56. Idris ZM, Chan CW, Kongere J, Hall T, Logedi J, Gitaka J, et al. Naturally acquired antibody response to Plasmodium falciparum describes heterogeneity in transmission on islands in Lake Victoria. Sci Rep. 2017;7(1):9123. Epub 20170822. doi: 10.1038/s41598-017-09585-4. PubMed PMID: 28831122; PubMed Central PMCID: PMCPMC5567232.

57. Balfour HH, Jr., Dunmire SK, Hogquist KA. Infectious mononucleosis. Clin Transl Immunology. 2015;4(2):e33. Epub 20150227. doi: 10.1038/cti.2015.1. PubMed PMID: 25774295; PubMed Central PMCID: PMCPMC4346501.

58. Guerrero-Ramos A, Patel M, Kadakia K, Haque T. Performance of the architect EBV antibody panel for determination of Epstein-Barr virus infection stage in immunocompetent adolescents and young adults with clinical suspicion of infectious mononucleosis. Clin Vaccine Immunol. 2014;21(6):817–23. Epub 20140402. doi: 10.1128/CVI.00754-13. PubMed PMID: 24695777; PubMed Central PMCID: PMCPMC4054247.

59. Lang D, Vornhagen R, Rothe M, Hinderer W, Sonneborn HH, Plachter B. Cross-reactivity of Epstein-Barr virus-specific immunoglobulin M antibodies with cytomegalovirus antigens containing glycine homopolymers. Clin Diagn Lab Immunol. 2001;8(4):747–56. doi: 10.1128/CDLI.8.4.747-756.2001. PubMed PMID: 11427421; PubMed Central PMCID: PMCPMC96137.

60. Berth M, Bosmans E. Acute parvovirus B19 infection frequently causes false-positive results in Epstein-Barr virus - and herpes simplex virus-specific immunoglobulin M determinations done on the Liaison platform. Clin Vaccine Immunol. 2009;16(3):372–5. Epub 20081230. doi: 10.1128/CVI.00380-08. PubMed PMID: 19116304; PubMed Central PMCID: PMCPMC2650871.

61. Tiguman GMB, Poll LB, Alves CEC, Pontes GS, Silva MT, Galvao TF. Seroprevalence of cytomegalovirus and its coinfection with Epstein-Barr virus in adult residents from Manaus: a population-based study. Rev Soc Bras Med Trop. 2020;53:e20190363. Epub 20200127. doi: 10.1590/0037-8682-0363-2019. PubMed PMID: 31994666; PubMed Central PMCID: PMCPMC7083370.

62. Nagata K, Kumata K, Nakayama Y, Satoh Y, Sugihara H, Hara S, et al. Epstein-Barr Virus Lytic Reactivation Activates B Cells Polyclonally and Induces Activation-Induced Cytidine Deaminase Expression: A Mechanism Underlying Autoimmunity and Its Contribution to Graves’ Disease. Viral Immunol. 2017;30(3):240–9. Epub 20170323. doi: 10.1089/vim.2016.0179. PubMed PMID: 28333576; PubMed Central PMCID: PMCPMC5393416.

63. Chen T, Song J, Liu H, Zheng H, Chen C. Positive Epstein-Barr virus detection in coronavirus disease 2019 (COVID-19) patients. Sci Rep. 2021;11(1):10902. Epub 20210525. doi: 10.1038/s41598-021-90351-y. PubMed PMID: 34035353; PubMed Central PMCID: PMCPMC8149409.

64. Smatti MK, Al-Sadeq DW, Ali NH, Pintus G, Abou-Saleh H, Nasrallah GK. Epstein-Barr Virus Epidemiology, Serology, and Genetic Variability of LMP-1 Oncogene Among Healthy Population: An Update. Front Oncol. 2018;8:211. Epub 20180613. doi: 10.3389/fonc.2018.00211. PubMed PMID: 29951372; PubMed Central PMCID: PMCPMC6008310.

65. Chabay P, Lens D, Hassan R, Rodriguez Pinilla SM, Valvert Gamboa F, Rivera I, et al. Lymphotropic Viruses EBV, KSHV and HTLV in Latin America: Epidemiology and Associated Malignancies. A Literature-Based Study by the RIAL-CYTED. Cancers (Basel). 2020;12(8). Epub 20200804. doi: 10.3390/cancers12082166. PubMed PMID: 32759793; PubMed Central PMCID: PMCPMC7464376.

66. Budiningsih I, Dachlan YP, Hadi U, Middeldorp JM. Quantitative cytokine level of TNF-alpha, IFN-gamma, IL-10, TGF-beta and circulating Epstein-Barr virus DNA load in individuals with acute Malaria due to P. falciparum or P. vivax or double infection in a Malaria endemic region in Indonesia. PLoS One. 2021;16(12):e0261923. Epub 20211228. doi: 10.1371/journal.pone.0261923. PubMed PMID: 34962938; PubMed Central PMCID: PMCPMC8714090.

67. Munz C. Modification of EBV-Associated Pathologies and Immune Control by Coinfections. Front Oncol. 2021;11:756480. Epub 20211028. doi: 10.3389/fonc.2021.756480. PubMed PMID: 34778072; PubMed Central PMCID: PMCPMC8581224.

